# ExCaPT: Explainable Cancer Prediction with Transformer-based models

**DOI:** 10.64898/2025.12.03.25341591

**Authors:** Ditsuhi Iskandaryan, Ferran Moratalla-Navarro, Alba Magallon-Baro, Lois Riobó-Mayo, Tomas Salas, Miguel Socolovsky, Victor Moreno

## Abstract

Cancer remains one of the most significant global health challenges. Despite advances in treatment, early detection remains a critical concern. The increasing availability of Electronic Health Records (EHR) offers a unique opportunity to enhance our understanding of patient health trajectories and develop more accurate risk prediction models. However, the complexity and heterogeneity of EHR data pose significant challenges for analysis and modeling. Over the years, a range of models, from traditional machine learning to advanced deep learning (DL) approaches, have been employed to address the multidimensional complexities of health data. Notably, transformer-based models have emerged as a promising solution for capturing longitudinal, sequential, and multimodal data. This work introduces ExCaPT, a transformer encoder-based predictive model designed to identify individuals at higher risk of developing colorectal cancer (CRC), while providing interpretable outputs. The model leverages a comprehensive dataset, incorporating features such as age, sex, smoking status, and longitudinal EHR data including disease and drug trajectories. ExCaPT had good performance in a test dataset with a ROC-AUC of 85.9 ± 0.1, 68.1 ± 0.3 sensitivity and 82.2 ± 0.1 specificity. These results outperform those of an LSTM model, used as reference for sequence data. This highlights the potential of transformer-based models in the early identification of high-risk cancer patients, marking an important step forward in the field of precision healthcare. Additionally, we employed several explainability approaches, including attention-based, embedding-based, and integrated gradients analyses, which allowed us to identify key input features, visualize latent representations, and quantify the contributions of different features to the predictions, providing complementary insights into the model’s decision-making process.

**Highlights:**

- ExCaPT, a transformer-encoder model using demographic, disease, and medication trajectories from EHR data, predicts early colorectal cancer (CRC) risk with high performance.
- The model provides interpretability through attention scores, embedding-based analyses, and integrated gradients to reveal influential features.
- This modeling framework is generalizable and can be extended to pre-diction for other cancer types.

**GRAPHICAL ABSTRACT:** 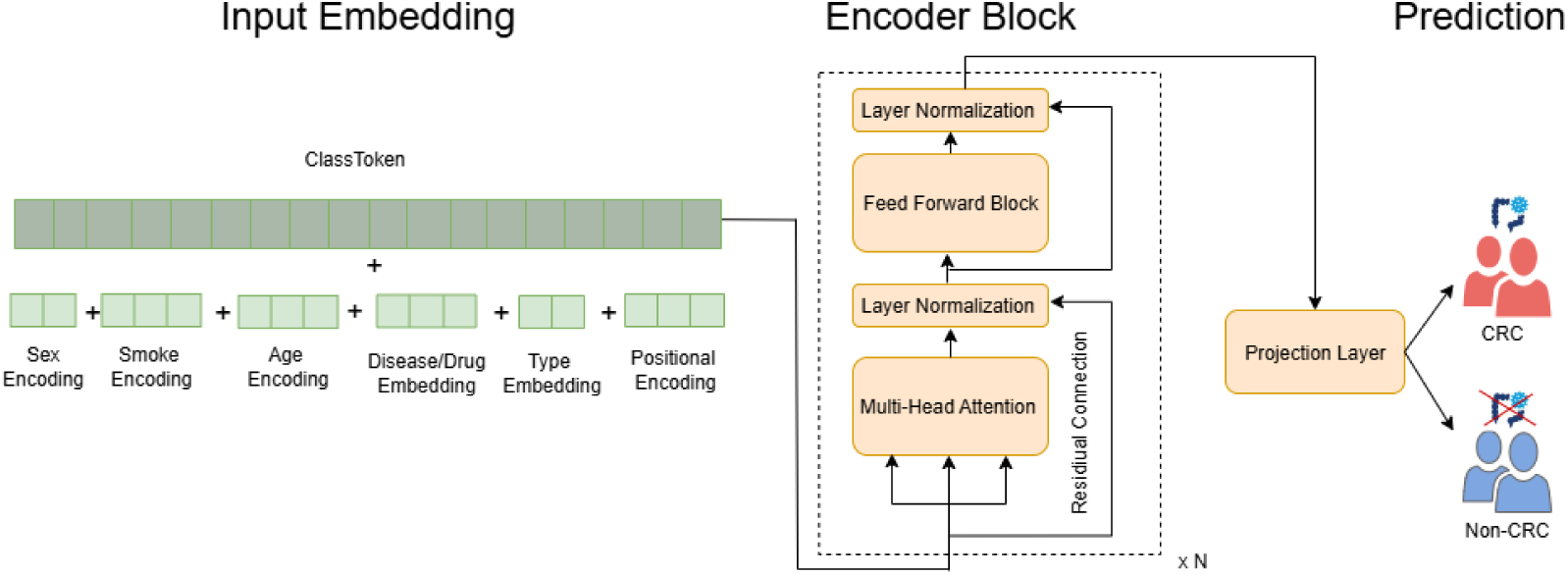

## 1. Introduction

Colorectal cancer (CRC), among the various types of cancer, stands out due to its high prevalence and mortality rates. It is the third most common cancer globally, following lung and breast cancer, and is the second leading cause of cancer-related deaths [1]. In 2020, approximately 1.9 million new cases of CRC were diagnosed globally, accounting for about 10% of all new cancer cases. That same year, CRC led to 930,000 deaths, representing 9.4% of all cancer-related deaths [1]. About 25-30% of CRC cases are linked to lifestyle factors such as diet, smoking, physical inactivity, and obesity. Additionally, 5-10% of cases result from inherited genetic conditions, including Lynch syndrome and familial adenomatous polyposis [2, 3].

Screening of CRC has proven to reduce mortality and is implemented in most developped countries either using a first test that detects occult blood in faeces using an immunochemichal reaction (FIT), followed by colonoscopy in those subjects that test possitive, or directly inviting to colonoscopy [4]. As of today, only age and familily history of CRC are used as stratification factors to define which subjects should be screened. One of the most promising ways to improve early CRC detection is through advanced predictive models that identify individuals at higher risk and should be invited to screening.

In addition to age and family medical history, the analysis of diverse data, such as demographic information, smoking history, previous disease history and prescription drug use could lead to the definition of a more personalized risk profile for patients. In recent years, artificial intelligence (AI) tools have become increasingly popular, especially in the medical field. By harnessing the power of these tools, healthcare professionals can provide quick, accurate solutions to critical issues, such as the early detection of cancer [5, 6].

Several key studies have contributed to this evolving field. For example, Gollangi et al. [7] aimed to evaluate various AI algorithms for classifying and predicting cancer outcomes using EHRs. Cooper et al. [8] focused on improving screening accuracy and identify individuals at higher risk of CRC by using primary care data. Morin et al. [9] introduced MEDomics, an AI system integrating multimodal health data for real-time cancer prognostication. The authors of the following work [10] focused on pancreatic cancer prediction by implemeting CancerRiskNet, which is based on GRU and Transformer. Zhou et al.[11] presented a transformer-based model fine-tuned on a cancer-specific corpus to enhance the extraction of breast cancer phenotypes from clinical texts. Yang et al. [12] proposed Transformer EHR, which is encoder-decoder based transformer aiming future diseases and outcomes of a patient using previous records. More recently, Shmatko et al. [13] have used a transformer with a modified GPT-2 architecture to analyzed the natural history of over 1000 diseases in the UK Biobank cohort, showing good predictive accuracy in the Danish population. Li et al. [14] focused on early detection of non-small cell lung cancer using EHR data. Table 1 lists the main features extracted from the reviewed literature.

**Table 1:** Features of the papers devoted to the disease prediction using ML and DL approaches.

| Work (Y) | Target/Goal | Methods | Data (N) | Metrics |
| --- | --- | --- | --- | --- |
| [7] (2020) | Breast cancer | GNB, CNNs, KNN, MobileNetV2 | CBIS-DDSM (5000) | F1-score, Sensitivity, Accuracy. |
| [8] (2020) | CRC | Cox + MFP | NHS-BCSP (292,059) | C-stat, D, calibration curve, R<sup>2</sup> |
| [9] (2021) | PCSP | MEDomics | Thousands |  |
| [11] (2022) | EBCP | CancerBERT | 21,291 | F1-Score |
| [10] (2023) | Pancreatic cancer | CancerRiskNet | DNPR, US-VA(6M,3M) | AUROC |
| [12] (2023) | PCP,ISHP | Transformehr | MIMIC-IV (6.5 M) | PPV,AUROC,AUPRC |
| [14] (2025) | NSCLC detection | Elastic-net+Cox | MGB-EHR (3546; 19040; 563,817 PY) | PPV,AUROC,Specificity |
| [13] (2025) | FDP | GPT-2 | UK Biobank (.4 M), Danish population (1.2 M) | AUC |

NSCLC-Non-Small Cell Lung Cancer, PCP-Pancreatic Cancer Prediction, EBCP-Extract Breast Cancer Phenotypes, BCSP-Bowel Cancer Screening Programme, ISHP-Intentional Self-Harm Prediction, FDP-Future Disease Prediction, PCSP-Pan-Cancer Survival Prognosis, DNPR-Danish National Patient Registry, US-VA-US Veterans Affairs, MGB-Mass General Brigham. Acc-Accuracy, PPV-Positive Predictive Value, PY-Patient-Years, MFP-Multivariable Fractional Polynomials.

In this study we introduce ExCaPT, a transformer-based predictive model for assessing CRC risk. The key feature of the transformer is the use of the self-attention mechanism [15], which allows the model to capture complex relationships between different parts of the input sequence. This mechanism enables each element (such as a feature or data point) to attend to all other elements in the sequence, making it effective for understanding dependencies, regardless of their position in the input. In the context of disease prediction, this ability allows the model to recognize patterns across various features (e.g., medical diagnosis and drugs used, patient demographics) that may be crucial for accurate predictions. Also, the model integrates explainability techniques to ensure transparency and interpretability in its decision-making process. Our aim is to develop a CRC risk prediction model that uses routinely available data to provide a better risk stratification and improve screening outcomes.

## 2. Methods

### 2.1. Ethics Statement

This study uses anonymized patient data. The Bellvitge Hospital Ethics Committee approved the study protocol (number PR320/18) waiving the need to obtain informed consent, given the large population analyzed and the anonymization procedure used by the data provider.

### 2.2. Patients

The dataset used in this work included primary care health data corresponding to the adult population of Catalonia, Spain, covered by the public health system (Catalan Health Institute), which is approximately 80% of the whole population. Data was provided anonimyzed by the Public Data Analysis for Health Research and Innovation Program (PADRIS: Programa d’analítica de dades per a la recerca i la innovació en salut) [16], managed by the Healthcare Quality and Evaluation Agency of Catalonia (AQuAS). To increase anonimity, patients in municipality strata with less than 10 subjects were not retrieved. The initial data extraction followed a case-control design, where all cancer cases diagnosed during the period Jan 2012 to Dec 2017 were identified, and for each cancer diagnosis, up to 10 controls were selected from the strata of the same sex, birth year (± 3 years) and municipality (∼ 54,000 strata). Since extracting information on drugs costly, only two controls per case were selected for extraction. Information on diseases and drugs for cases and controls were right censored at one year before the age of the diagnosis of the index cancer in the case (cases and controls could have other previous cancers as diseases). For this analysis, only CRC cases and their matched controls were selected, following the procedure described in the flowchart of Figure 1. Stage 1 presents the raw number of subjects extracted in the case and control groups. Stages 2 to 5 involved a series of data cleaning steps such as excluding patients who only had diseases out of the study period and removing subjects with no prior diseases or only one disease but with low prevalence. Subsequently, we filtered and selected CRC cases along with their matched controls. Each case was matched to at least one control. For training, we used a balanced dataset, pairing each case with a randomly selected control. For testing, we used the full set of available controls and their corresponding cases without any additional adjustments, and we also included the set of controls that had been excluded during the training-stage balancing process.

**Figure 1.**
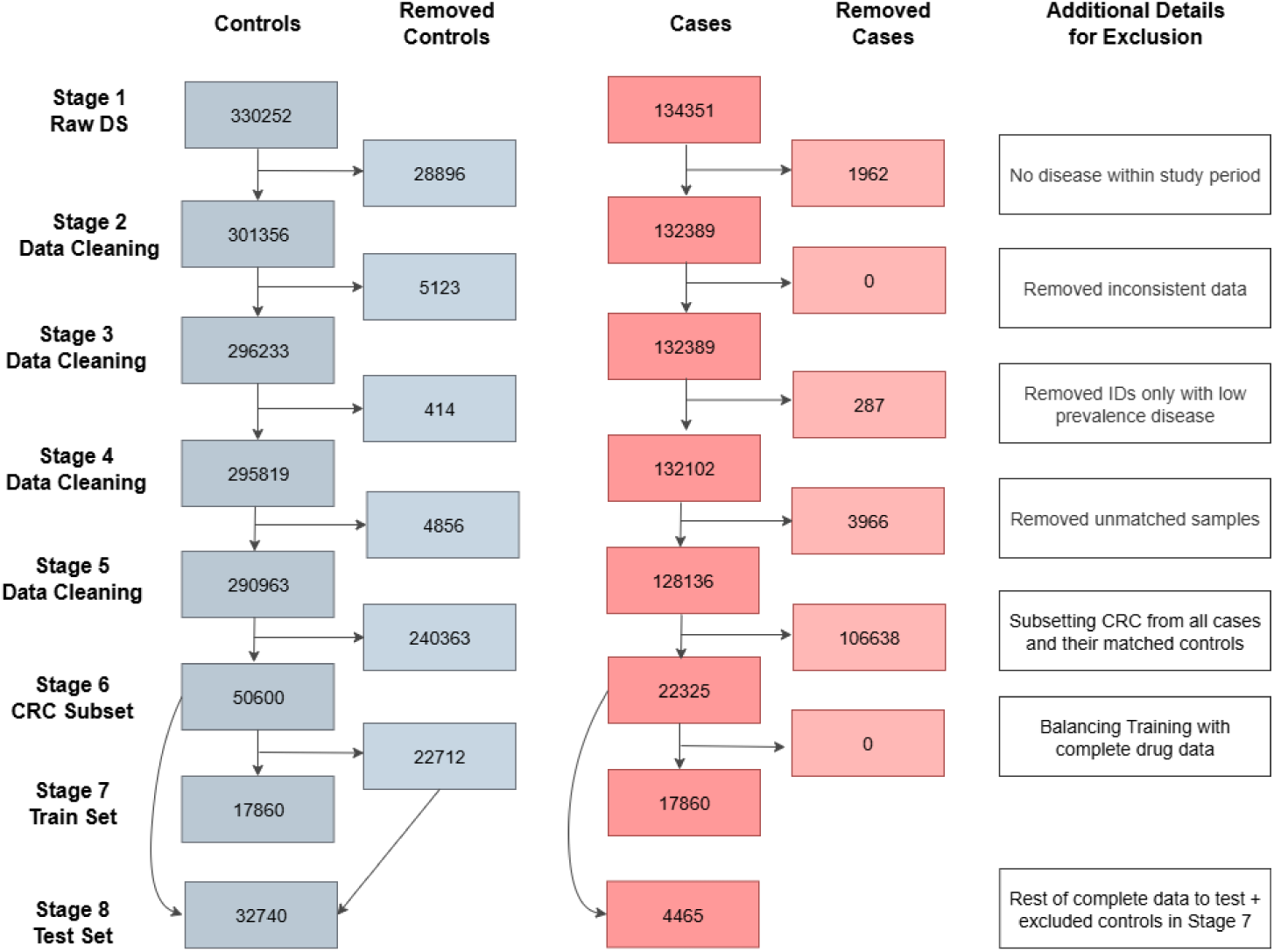
Overview of the cohort selection process, including inclusion and exclusion criteria across each stage.

### 2.3. Data Preprocessing

Diseases were coded using the International Classification of Diseases, Tenth Revision (ICD-10), grouped at the 3-character level, which captures general disease categories (e.g., E11 for type 2 diabetes mellitus). Prescribed drugs were mapped using the Anatomical Therapeutic Chemical (ATC) classification. Drug prescriptions were grouped at the 5-character level of the ATC classification system (e.g., A10BB for sulfonylureas), providing a detailed view of pharmacological treatments. Several ICD-10 codes were grouped together based on their clinical similarity.

Only drugs and diseases that were commonly observed across patients were retained, excluding those occurring in less than 1,000 patients in the complete dataset. This approach allowed us to exclude rare items from the analysis.

To incorporate both diseases and drugs in a unified input format, we combined them and a special token [SEP] was inserted to clearly separate disease codes from drug codes. This helped the model distinguish between the two modalities within the same sequence. Additionally, we constructed a parallel binary indicator vector where entries corresponding to diseases were marked as 0 and those corresponding to drugs as 1. This auxiliary vector was included as an additional input to help the model differentiate between the types of medical events.

Sex, age (of CRC diagnosis for cases and the same for their matched controls) were available for all individuals. Smoking status, being a relevant risk factor for multiple diseases including CRC, was also extracted for most of subjects.

Since each patient had a different length of event data, we standardized these lengths for effective input into the transformer model. To do this, we calculated the maximum event length across all patients in the case cohort, ensuring that we captured all relevant information leading up to the onset of colorectal cancer. This maximum length was then applied to both the cases and the controls. While some control samples may exceed this length slightly, these few instances will have minimal impact on the overall dataset.

The next step was to balance the cases and controls in the training set across all features. While the distributions of age and sex were balanced between the two groups, smoking status remained imbalanced, as shown in Table 5. To address this, we adjusted the distribution to ensure equal representation of each smoking status category across both cohorts. We calculated the absolute differences in the number of patients for each smoking category between the cases and controls in the training set (see Table 2). Based on these differences, we increased the number of missing values in the control cohort and reduced the number of patients in the overrepresented categories to achieve balance.

**Table 2:** Number of patients by smoking status for cases and controls in the training set, with the absolute difference between cohorts.

| Smoking Status | Cases | Controls | Absolute Difference |
| --- | --- | --- | --- |
| Never | 6740 | 9027 | 2287 |
| Former | 1736 | 2209 | 473 |
| Current | 1350 | 2042 | 692 |
| Missing | 8034 | 4582 | 3452 |

The dataset was divided into training and test sets. We first split the cases using an 80/20 ratio. In the training set, cases and controls were paired in a 1:1 ratio (50% cases and 50% controls). This balanced dataset was crucial for preventing any class imbalance that could skew model performance. The test set maintained the same proportion of paired samples and additionally included the controls that had been excluded during the training-stage balancing process.

After balancing the dataset, the next step was converting the raw data into tokens, which was essential to feed the data into a transformer model. Table 3 summarizes the tokenization details.

**Table 3:** Details about tokenization results.

| Features | # Tokens | Tokens |
| --- | --- | --- |
| Sex | 2 | [0, 1] |
| Smoking Status | 4 | [0, 1, 2, 3] |
| Age | 120 | [0, 1, 2..., 119] |
| Diseases | 485 | [0, 1, 2,..., 484] |
| Drugs | 300 | [485, 486, ..., 784] |
| Special Tokens (PAD, MASK, CLS, SEP) | 4 | [785, 786, 787, 788] |

### 2.4. Model Architecture

The method used in this work was based on the Transformer architecture. A detailed explanation of the Transformer can be found in the paper by Vaswani *et al.* (2017) [15]. The Transformer consists of both encoder and decoder components, but for the purpose of this study, we focus on utilizing only the Transformer encoder.

The proposed model was designed to process input sequences and convert them into high-level representations, referred to as embeddings. These embeddings capture the relevant features of the input data and were used for disease prediction. Figure 2 presents the workflow of the proposed methodology, that consists of the following components: *Input Embedding*, *Encoder Block*, and *Prediction*.

**Figure 2:**
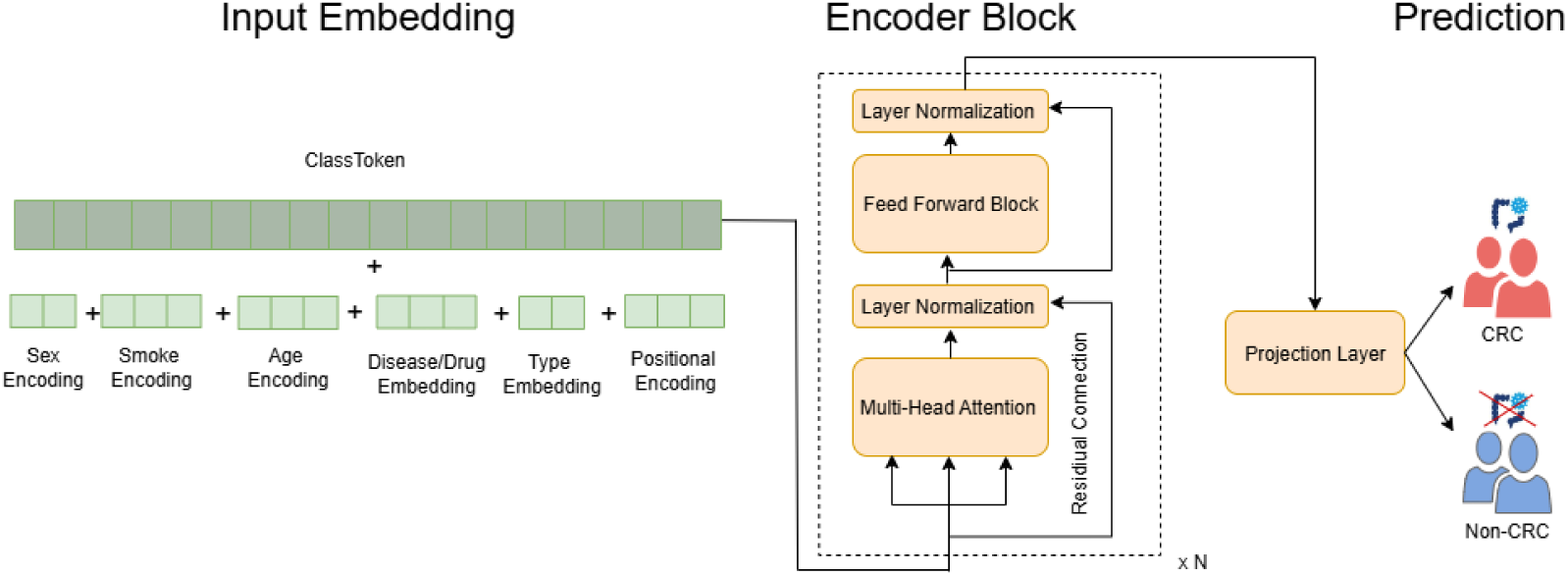
Workflow for CRC vs. Non-CRC prediction using a transformer encoder-based approach. The model comprises three main modules: Input Embedding, Encoder Block, and Prediction.

Input Embedding: this module maps heterogeneous clinical features into continuous vector spaces. It includes *Sex Encoding* (embeds sex-related information), *Smoke Encoding* (embeds smoking status), *Age Encoding* (embeds age-related information using a sinusoidal pattern), *Disease/Drug Embedding* (embeds disease-related and drug-related vocabularies), *Type Embedding* (binary vector where 0 indicates a disease and 1 indicates a drug), *Positional Encoding* (applies positional encoding to represent the position of each item in the input data, ensuring the model understands the sequential order), and *Class Token* (allows the model to learn a global representation for classification tasks).

Encoder Block: this module consists of *Multi-Head Attention* (allows the model to weigh the importance of each input feature in relation to others), *Feed-Forward Block* (helps the model learn complex, non-linear mappings from the attention output to the final representation), *Layer Normalization* (stabilizes training), and *Residual Connection* (aids in the training of deep networks and mitigates issues such as vanishing/exploding gradients).

Prediction: this module includes *Projection Layer*, which the model uses to project the output to make predictions.

### 2.5. Model Training

The Adam optimizer [17] was employed for weight updates. The loss function was based on BCE With Logits Loss which combines sigmoid activation and binary cross-entropy loss. We used gradient accumulation to increase the effective batch size, and a Cyclic Learning Rate (CyclicLR) scheduler.

### 2.6. Htpermarameter tuning

Several key hyperparameters were tuned to optimize the performance of the model. Table 4 provides a detailed description of hyperparameters, and the selected options are highlighted in bold.

**Table 4:**
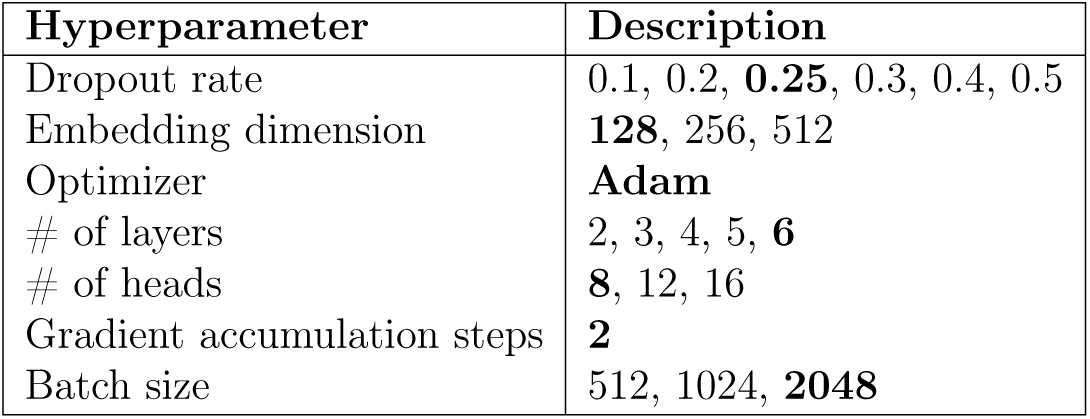
Description of the hyperparameters of the proposed model, with the selected options highlighted in bold.

### 2.7. Evaluation Metrics

Model performance was evaluated by the following evaluation metrics, namely precision, sensitivity, specificity, F1-Macro, Kappa value (*k* ), Mathew Correlation, balanced accuracy and AUC-ROC [18, 19, 20].

### 2.8. Attention-based analysis

To quantify the differences in attention weights between groups, we computed the Signal-to-Noise Ratio (SNR) for each disease-related element. The SNR was calculated as the difference in mean weights between CRC and non-CRC groups, normalized by the combined variability of both groups (Eq. 1):

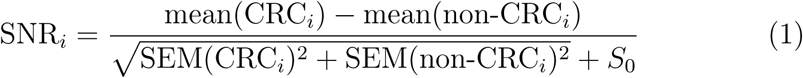

where mean(CRC*_i_*) and mean(non-CRC*_i_*) are the mean weights for the CRC and non-CRC groups, respectively, for element *i*, and SEM(CRC*_i_*) and SEM(non-CRC*_i_*) are the standard errors of the mean (SEM) for the CRC and non-CRC groups. To prevent division by very small denominators, a small constant *S*_0_ was added to the denominator (Eq. 2):

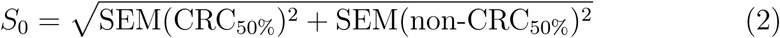

where SEM(CRC_50%_) and SEM(non-CRC_50%_) correspond to the median (50th percentile) standard errors of the mean across all CRC and non-CRC groups, respectively.

The SNR provides a measure of how strongly each disease and drug contributes to distinguishing between the two groups.

The analyses were carried out in a controlled Python 3.9 environment, using the Pytorch 1.10.1 DL library to build and train our models. In terms of hardware, we used two NVIDIA GPUs, Geforece RTX 3090, with 24 GB each.

## 3. Results

The obtained dataset has over 130,000 patients corresponding to all the cancer diagnoses in Catalonia, Spain, during the period Januray 2012 to December 2017. For each cancer case, 2 controls were selected, matched by birth year, sex, and municipality of residence. Data for controls were censored after the date of cancer diagnosis of the reference case. For this study, we analyzed the subset of 22,325 CRC cases and their 50,600 matched controls that were divided in a balanced train set of 17,860 CRC and controls and a validation set with the remaining subjects (Table 5).

Regarding features, we incorporated demographic data, including *age*, *sex*, and *smoking status*, with longitudinal EHR data including *disease* and *drug* trajectories. Table 5 shows patient characteristics.

### 3.1. Model development and validation

To assess the performance of the proposed model, we compared it against a well-established baseline, the Long Short-Term Memory (LSTM) network, a type of recurrent neural network (RNN) that is particularly effective for tasks involving sequential data, as they are designed to capture long-range dependencies within the data.

The performance of the model was evaluated using bootstrapping and a 30-iteration validation process. In each iteration, a bootstrapped subset of the test dataset was created to assess model stability. The model was trained, and metrics were calculated. Afterward, the mean values and 95% confidence intervals were computed to offer a more reliable measure of model performance. The results are presented in Figure 3 and Table 6.

**Figure 3:**
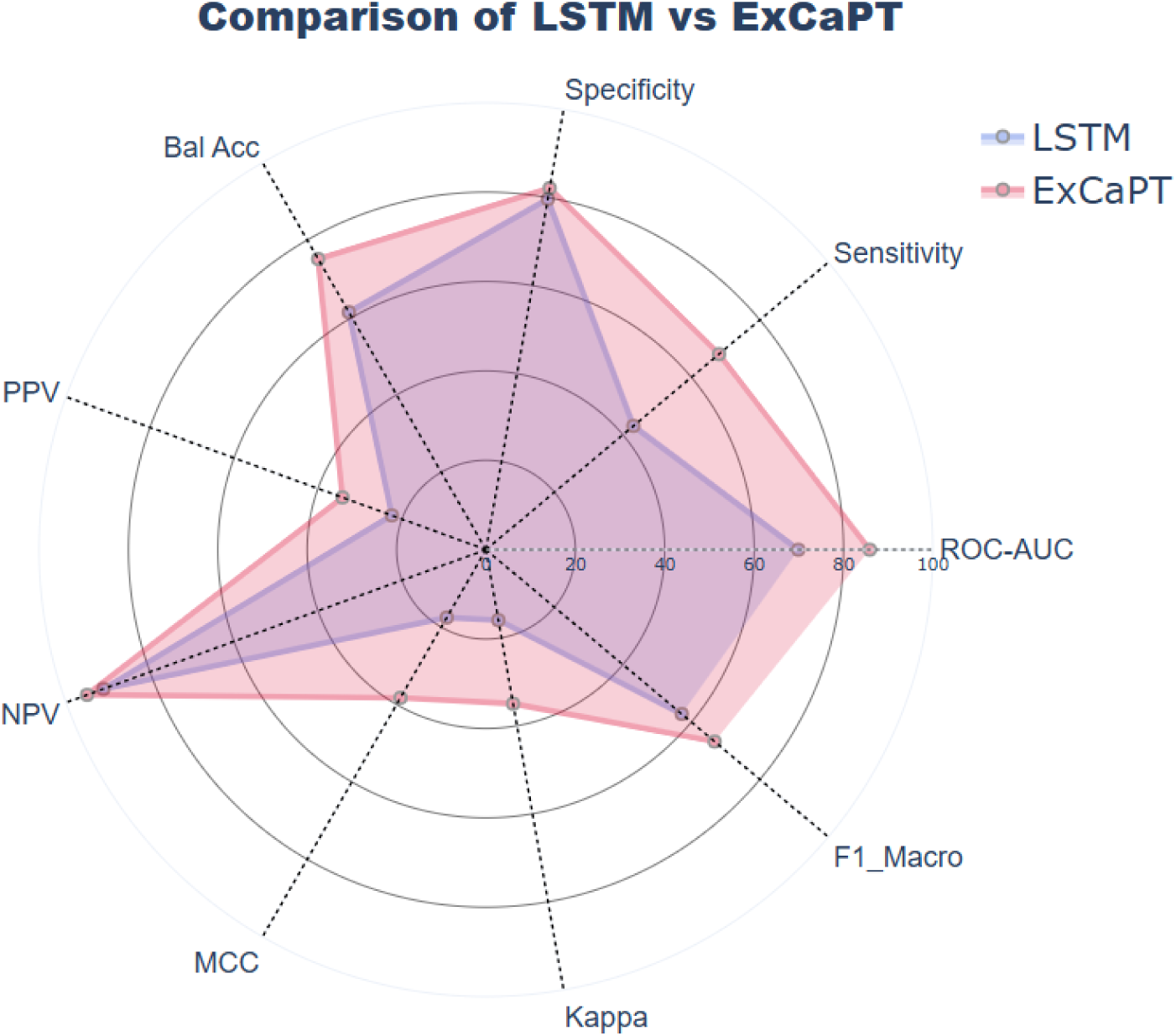
Evaluation metrics of LSTM and ExCaPT (all metrics in %, MCC and Kappa ×100 for scale). Bal. Acc.: Balanced accuracy; ROC-AUC: area under the Receiver-Operating Characteristic curve; MCC: Mathews correlation coefficient; PPV: Positive Predictive Value (Precision); NPV: Negative Predictive Value. The cutoff probability was 0.5, and the test dataset had a 12% prevalence of CRC.

**Table 5:** Descriptive analysis of dataset.

| Characteristics | Groups | Train Set | Test Set |
| --- | --- | --- | --- |
| Number of patients | Cases | 17860 | 4465 |
|  | Controls | 17860 | 32740 |
| **Sex** |  |  |  |
| Female | Cases | 7828 | 1951 |
|  | Controls | 7828 | 14675 |
| Male | Cases | 10032 | 2514 |
|  | Controls | 10032 | 18065 |
| **Age** |  |  |  |
| Mean (SD) | Cases | 66.6 (11.9) | 66.6 (11.9) |
|  | Controls | 63.6 (12.3) | 63.04 (11.9) |
| < 50 | Cases | 781 | 205 |
|  | Controls | 781 | 1763 |
| 50 – 70 | Cases | 8221 | 2050 |
|  | Controls | 8221 | 16362 |
| > 70 | Cases | 8858 | 2210 |
|  | Controls | 8858 | 14615 |
| **Smoke** |  |  |  |
| Never | Cases | 6740 | 1730 |
|  | Controls | 9027 | 16237 |
| Former | Cases | 1736 | 452 |
|  | Controls | 2209 | 4125 |
| Current | Cases | 1350 | 330 |
|  | Controls | 2042 | 3550 |
| Missing | Cases | 8034 | 1953 |
|  | Controls | 4582 | 8828 |
| **Disease** |  |  |  |
| Number of unique diseases | 485 |  |  |
| Mean (SD) of Length | Cases | 10.4 (7.3) | 10.4 (7.2) |
|  | Controls | 12.2 (7.0) | 15.2 (9.1) |
| **Drugs** |  |  |  |
| Number of unique drugs | 300 |  |  |
| Mean (SD) of Length | Cases | 7.0 (6.1) | 7.0 (5.9) |
|  | Controls | 7.5 (5.8) | 8.8 (6.6) |

**Table 6:** Evaluation metrics for ExCaPT, stratified by age group, sex, and input sequence length, and LSTM (*ExCaPT, the proposed method, denotes evaluation on the full population with sequence length ≥ 1).

| Group | ROC-AUC | Sensitivity | Specificity | Bal. Acc. | PPV | NPV | MCC | Kappa | F1-Macro |
| --- | --- | --- | --- | --- | --- | --- | --- | --- | --- |
| ExCaPT* | 85.9 ± 0.1 | 68.1 ± 0.3 | 82.2 ± 0.1 | 75.1 ± 0.1 | 34.2 ± 0.2 | 94.98 ± 0.1 | 0.38 ± 0.002 | 0.35 ± 0.002 | 66.8 ± 0.1 |
| **Age Groups** |  |  |  |  |  |  |  |  |  |
| < 50 | 73.5 ± 0.8 | 67.0 ± 1.3 | 66.5 ± 0.4 | 66.8 ± 0.7 | 18.7 ± 0.6 | 94.6 ± 0.2 | 0.21 ± 0.01 | 0.16 ± 0.008 | 53.6 ± 0.5 |
| 50-70 | 82.7 ± 0.1 | 62.2 ± 0.4 | 81.1 ± 0.1 | 71.6 ± 0.2 | 29.2 ± 0.3 | 94.5 ± 0.1 | 0.32 ± 0.003 | 0.29 ± 0.003 | 63.5 ± 0.1 |
| > 70 | 90.03 ± 0.1 | 73.9 ± 0.4 | 85.5 ± 0.1 | 79.7 ± 0.2 | 43.5 ± 0.3 | 95.6 ± 0.1 | 0.48 ± 0.003 | 0.46 ± 0.003 | 72.5 ± 0.2 |
| **Disease Sequence Length for patients under 50** |  |  |  |  |  |  |  |  |  |
| $\geq 2$ | 72.9 ± 0.7 | 67.4 ± 1.3 | 65.5 ± 0.5 | 66.5 ± 0.7 | 16.8 ± 0.6 | 95.1 ± 0.3 | 0.2 ± 0.008 | 0.14 ± 0.007 | 52.2 ± 0.5 |
| $\geq 3$ | 72.9 ± 0.7 | 66.4 ± 1.3 | 65.0 ± 0.5 | 65.7 ± 0.7 | 16.4 ± 0.6 | 94.95 ± 0.3 | 0.19 ± 0.009 | 0.13 ± 0.007 | 51.7 ± 0.5 |
| **Sex** |  |  |  |  |  |  |  |  |  |
| Male | 84.03 ± 0.1 | 66.1 ± 0.4 | 80.5 ± 0.1 | 73.3 ± 0.2 | 31.9 ± 0.2 | 94.5 ± 0.1 | 0.35 ± 0.003 | 0.32 ± 0.003 | 64.99 ± 0.2 |
| Female | 87.9 ± 0.2 | 70.0 ± 0.4 | 84.4 ± 0.1 | 77.2 ± 0.2 | 37.3 ± 0.3 | 95.5 ± 0.1 | 0.42 ± 0.004 | 0.39 ± 0.004 | 69.1 ± 0.2 |
| **Disease Sequence Length** |  |  |  |  |  |  |  |  |  |
| $\geq 2$ | 85.8 ± 0.1 | 65.3 ± 0.2 | 83.5 ± 0.1 | 74.4 ± 0.1 | 32.5 ± 0.2 | 95.2 ± 0.04 | 0.37 ± 0.002 | 0.34 ± 0.002 | 66.2 ± 0.1 |
| $\geq 3$ | 86.3 ± 0.1 | 65.4 ± 0.3 | 84.1 ± 0.1 | 74.7 ± 0.1 | 33.2 ± 0.1 | 95.2 ± 0.1 | 0.38 ± 0.002 | 0.35 ± 0.002 | 66.7 ± 0.1 |
| **LSTM** | 69.9 ± 0.2 | 43.1 ± 0.3 | 79.7 ± 0.1 | 61.4 ± 0.2 | 22.4 ± 0.2 | 91.1 ± 0.1 | 0.18 ± 0.003 | 0.16 ± 0.003 | 57.2 ± 0.2 |

Figure 4a presents the confusion matrix for the test set, illustrating the distribution of predicted versus actual labels. We used as cutoff the 0.5 pre-dicted probability of CRC, though this threshold could be increased in practice to reduce the number of diagnostic tests required to confirm the diagnosis or reduced if higher sensitivity is desired. Regarding positive predictve value (PPV, precision), the test dataset had higher prevalence (11%) than the real population, where CRC only occurs in less than 3%. Figure 5 shows how metrics would vary with diverse cutoffs and the real CRC prevalence in the population.

**Figure 4:**
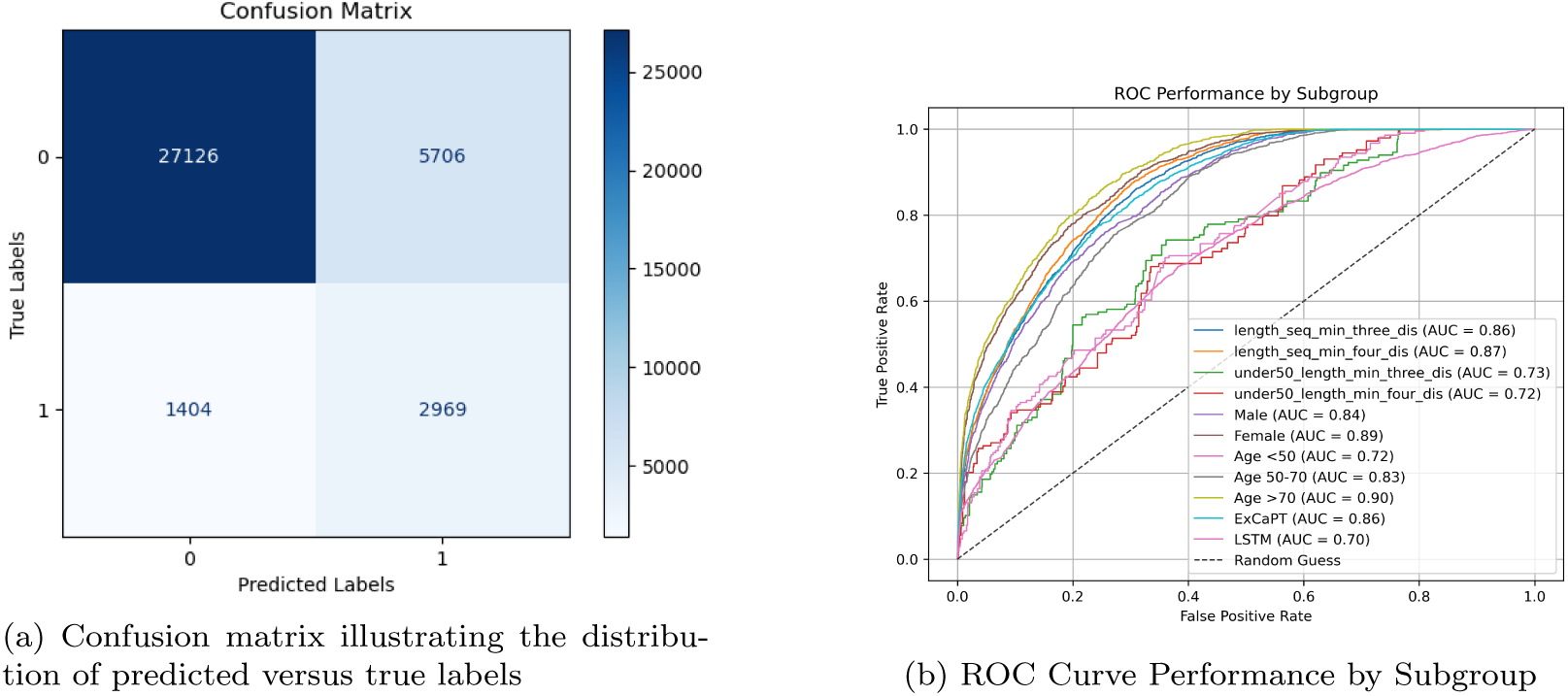
(a) Confusion matrix illustrating the distribution of predicted (CRC probability *>* 0.5) vs. true labels. (b) ROC Curve Performance by Subgroups (*length_seq_min_three_dis*: sequences with a minimum of three disease events; *length_seq_min_four_dis*: sequences with a minimum of four disease events; *un-der50_length_min_three_dis*: sequences under age 50 containig at least three disease; *under50_length_min_four_dis*: sequences under age 50 containig at least four disease; *Male*: male patients; *Female*: female patients; *Age&lt;50*: patients aged 50; *Age 50-70*: patients aged 50–70; *Age >70*: patients aged 70+; ExCaPT and LSTM).

**Figure 5:**
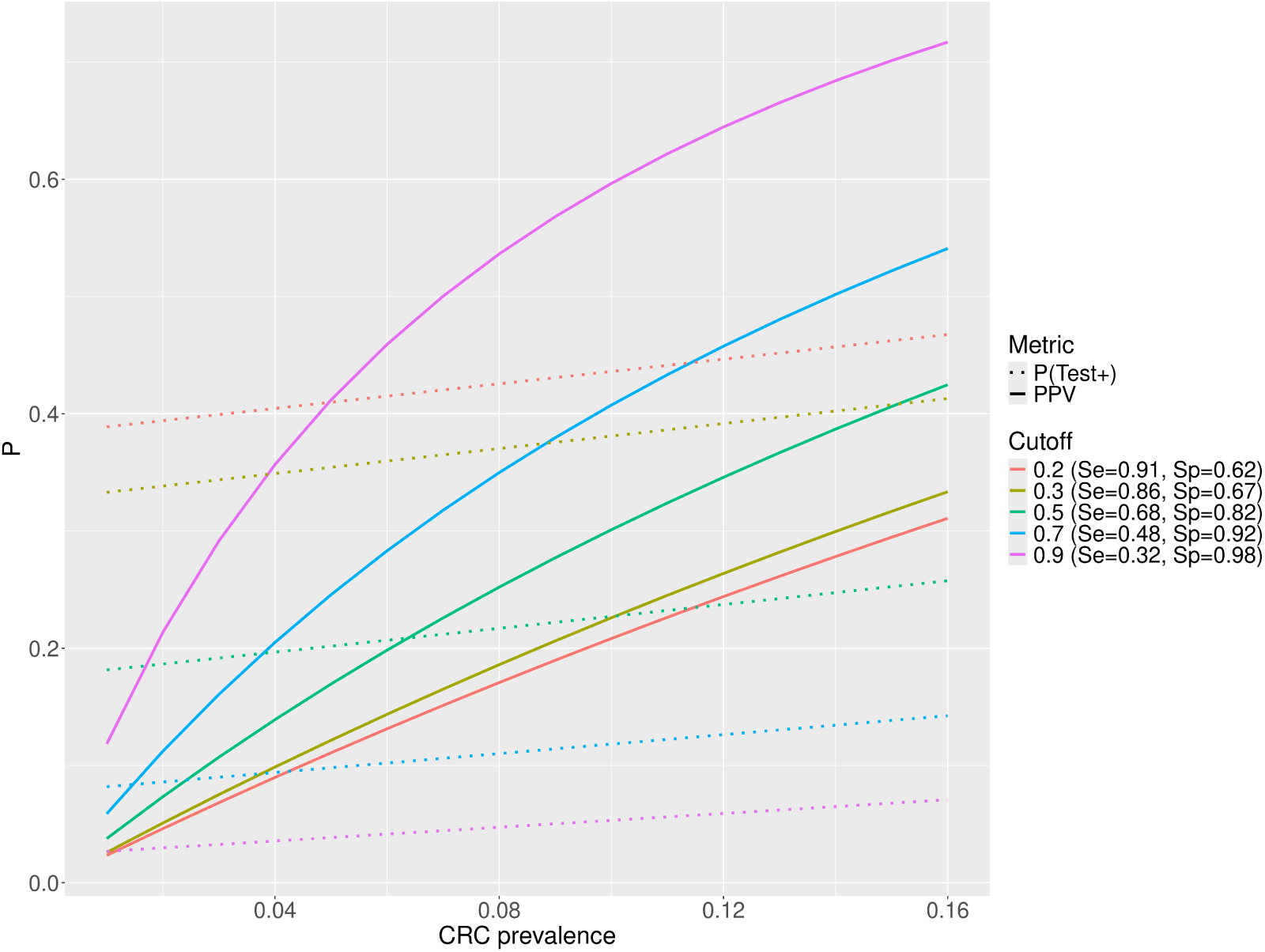
Model performance metrics across different cutoff thresholds.

Furthermore, we conducted a stratified evaluation of ExCaPT to assess its generalizability and effectiveness across diverse demographic and clinical subgroups. This included analysis by age group (< 50 years, 50–70 years, and > 70 years), sex (male and female), and disease sequence length (patients with at least 1, 2, or 3 recorded diagnoses). For patients under the age of 50, we also examined subsets of patients with at least 2 or 3 diseases. Additionally, we performed experiments in which the lengths of disease and drug sequences were randomly equalized to analyze the model’s robustness to input variability. The results of this stratified analysis are presented in Table 6. Figure 4b displays the ROC curves for the corresponding subgroups.

Predictive accuracy was high in all subgroups. Since CRC is less frequent among young people, the high accuracy observed even in the age *<* 50 might be related to the model learning that few diagnosis increased the likelihood of being in the CRC group. This was confirmed when we analysed models subsetting patients and controls with at least 2 or 3 diagnoses, obtaining lower accuracy metrics. The accuracy also decreased when we sampled diagnosis from controls to equalise the sequence length, indicating that the number of available diagnosis was informative.

### 3.2. Explainability

In this subsection, we focus on the explainability of the model’s performance, to gain deeper insights into the decision-making process of the model. We employed the following approaches: attention-based analysis, embedding-based analysis, and integrated gradients-based analysis.

#### 3.2.1. Attention-based Analysis

A key contribution of this work is the integration of the attention mechanism, which significantly enhances model explainability by revealing the areas of focus during predictions. To investigate the decision-making process, we visualised the attention weights associated with the class token, which generalises the attention across all diseases and drugs. This allowed us identifying which diseases and drugs had the most influence on cancer prediction.

These visualizations were generated by averaging the attention weights across all attention heads in the last layer. We then aggregated attention maps across samples and computed the mean and SEM, providing a comprehensive view of attention distribution while accounting for variability and uncertainty.

Differences between groups were estimated by SNR of attention weights for each disease-related element. The SNR provides a measure of how strongly each disease and drug contributes to distinguishing between the two groups. We annotated some codes (see Table 7) based on their relevance as risk factors for CRC (see Figure 6). Detailed information, including the mean, SEM, and SNR of class token attention weights for all tokens, is provided in Supplementary Table S1.

**Figure 6.**
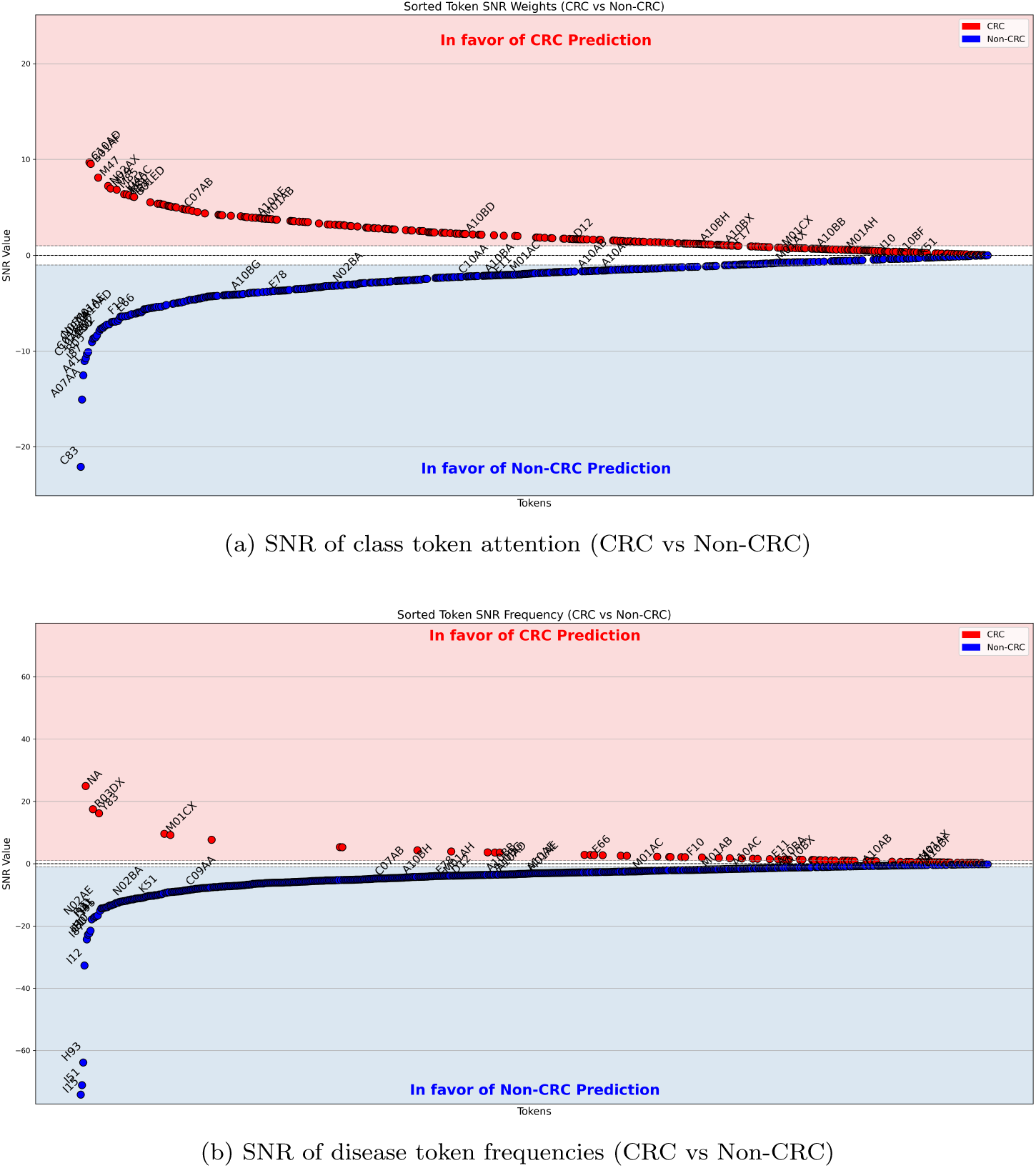
Attention-based explainability: (a) SNR analysis for class token attention (CRC vs Non-CRC), (b) SNR analysis of disease token frequencies (CRC vs Non-CRC).

**Table 7:** ICD-10 and ATC Codes with Descriptions.

| ICD-10 Code | Description | ATC Code | Description |
| --- | --- | --- | --- |
| D12 | Benign neoplasm of the colorectum | A10 | Antidiabetics |
| E11 | Type 2 diabetes mellitus | C07AB | Beta-blockers |
| E66 | Obesity | C09AA | ACE inhibitors |
| E78 | Lipidemias | C10AA | Statins |
| F10 | Alcohol abuse | M01 | Non-Steroidal Anti-inflammatory |
| F17 | Tobacco addiction | N02BA | Aspirin |
| I10 | Hypertension |  |  |
| K50 | Crohn’s disease |  |  |
| K51 | Ulcerative colitis |  |  |

We further calculated the SNR of disease frequencies using bootstrap sampling to assess whether the attention weights correlated with disease frequencies. The results are shown in Figure 6. Complete information, including the frequency of cases and controls, odds ratio, and 95% CI, is provided in Supplementary Tables S2 and S3.

#### 3.2.2. Embedding-based Analysis

In addition to attention weights, we extract embeddings from the model and projected them into a 2D space using t-SNE based on different features to explore potential patterns or clustering in the embedding space (Figure 7).

**Figure 7.**
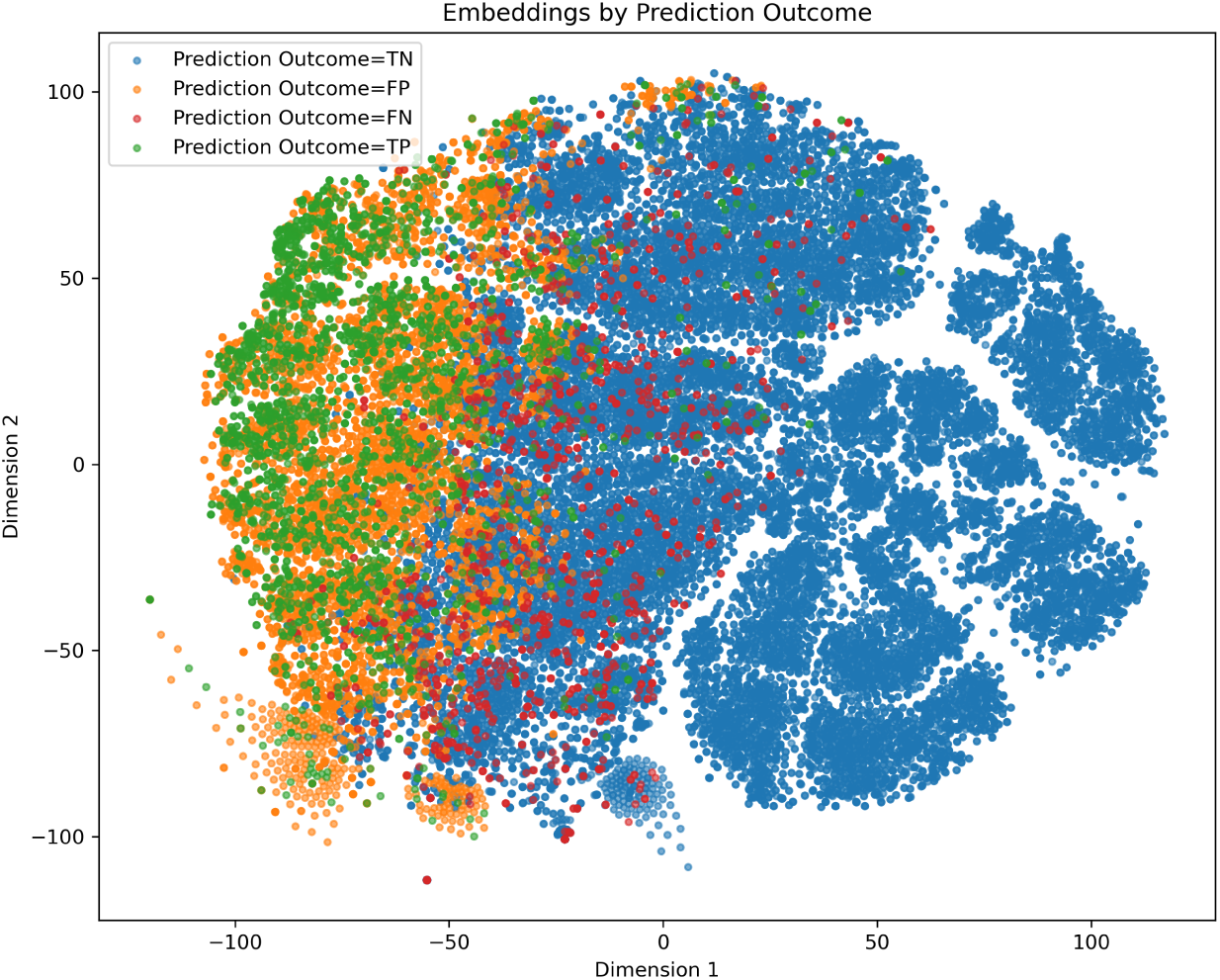
t-SNE visualization of model embeddings. Colors correspond to true positives (TP), true negatives (TN), false positives (FP), and false negatives (FN), revealing patterns in how the model separates correct and incorrect predictions in the embedding space.

#### 3.2.3. Integrated Gradients Attribution Analysis

To interpret model predictions, we applied Integrated Gradients (IG) [21] to quantify the contribution of input tokens to the model’s decisions. Specifically, we focused on correctly predicted samples belonging to the positive class (i.e., target class = 1). For each such instance in the test set, we computed IG with respect to the input disease-drug token embeddings.

The attributions were computed by interpolating between a baseline (all tokens replaced with the padding token) and the actual input embeddings. The attribution model wraps the encoder and projection layer of the model, using the output corresponding to the [CLS] token to estimate prediction scores. Additional features (sex, smoking status, age, and drug type) were provided as static arguments to the attribution function. We used 200 interpolation steps for the IG computation.

Optionally, to enable token-level analysis across the dataset, the attributions were aggregated over all matching samples. For each token (excluding padding), we calculated both the mean attribution score and SEM to assess the overall importance of each token. Figure 8 shows the result. Detailed information, including the mean, SEM, and SNR of IG for all tokens, is provided in Supplementary Table S4.

**Figure 8:**
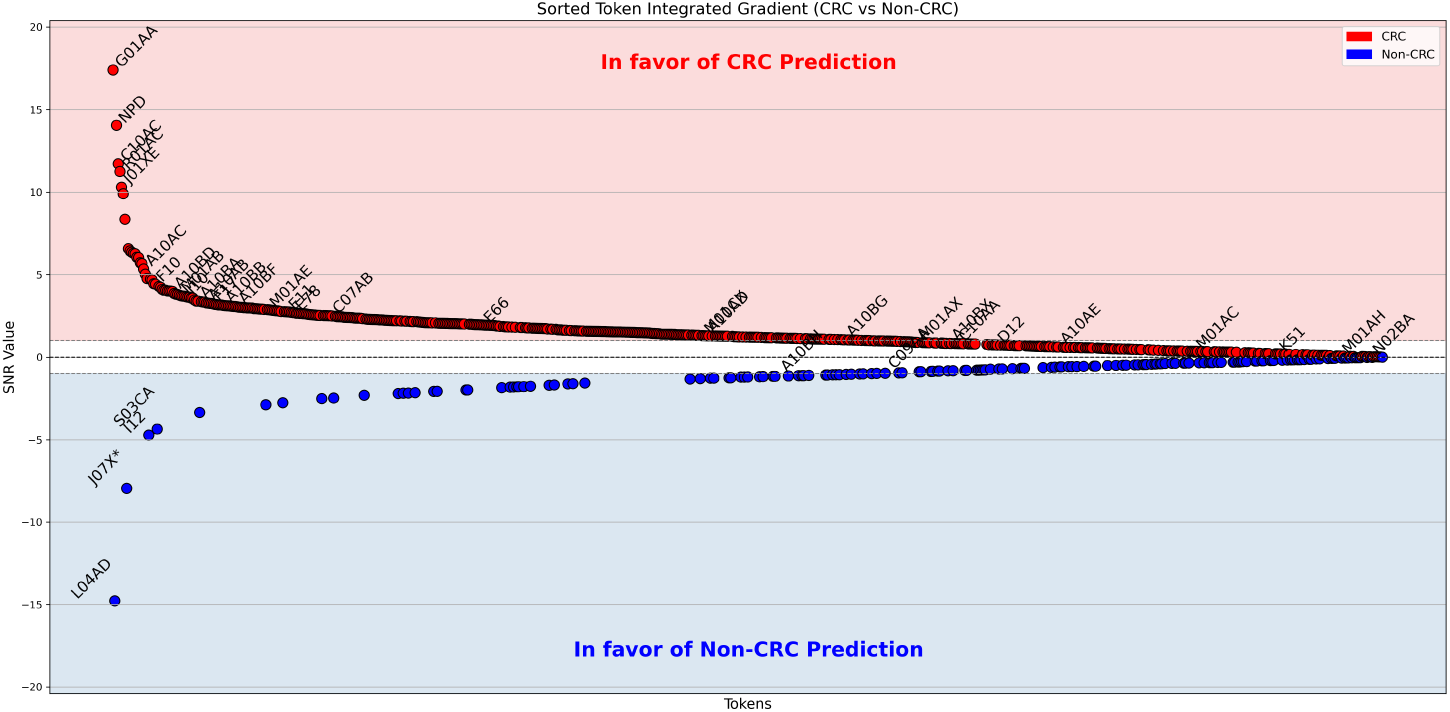
SNR of of Integrated Gradients Attribution Scores.

### 3.3. Ablation Analysis

Additionally, an ablation study was conducted to systematically evaluate the contribution of individual model components and input features, enabling us to identify the most critical factors driving model performance. We focused on the following components: disease and drug embedding, sex embedding, smoke embedding, disease and drug type embedding, age embedding, positional encoding, and class token embedding. Table 8 presents the results for ten different configurations: 1. *Full Model* (all components enabled); 2. *All Disabled* (all components disabled); 3. *Only Class Token Enabled* (only class token embedding enabled); 4. *No Class Token* (all components enabled except class token embedding disabled). 5. *No Sex Embedding* (sex embedding disabled); 6. *No Smoke Embedding* (smoke embedding disabled); 7. *No Age Embedding* (age embedding disabled); 8. *No Positional Encoding* (positional encoding disabled); 9. *No Age & Positional Encoding* (age embedding and positional encoding disabled); 10. *No Disease & Drug Embedding* (disease and drug embedding disabled).

**Table 8:** Ablation study results summarizing evaluation metrics with confidence intervals for the proposed model under various configuration settings.

| Methods | ROC-AUC | Sensitivity | Specificity | Bal. Acc. | PPV | NPV | MCC | Kappa | F1-Macro |
| --- | --- | --- | --- | --- | --- | --- | --- | --- | --- |
| Full Model | 85.9 ± 0.1 | 68.1 ± 0.3 | 82.2 ± 0.1 | 75.1 ± 0.1 | 34.2 ± 0.2 | 94.98 ± 0.1 | 0.38 ± 0.002 | 0.35 ± 0.002 | 66.8 ± 0.1 |
| Only Class Token Enabled | 61.5 ± 0.2 | 27.7 ± 0.2 | 83.4 ± 0.1 | 55.6 ± 0.1 | 18.6 ± 0.2 | 89.4 ± 0.1 | 0.1 ± 0.002 | 0.09 ± 0.002 | 54.3 ± 0.1 |
| No Class Token | 84.5 ± 0.1 | 43.7 ± 0.2 | 92.6 ± 0.1 | 68.2 ± 0.1 | 44.7 ± 0.3 | 92.3 ± 0.1 | 0.37 ± 0.002 | 0.37 ± 0.002 | 68.3 ± 0.1 |
| No Sex Embedding | 85.8 ± 0.1 | 62.5 ± 0.3 | 84.95 ± 0.1 | 73.7 ± 0.2 | 36.1 ± 0.3 | 94.3 ± 0.1 | 0.38 ± 0.003 | 0.36 ± 0.003 | 67.6 ± 0.1 |
| No Smoke Embedding | 83.9 ± 0.1 | 62.6 ± 0.2 | 81.4 ± 0.1 | 71.97 ± 0.1 | 31.5 ± 0.2 | 94.1 ± 0.1 | 0.34 ± 0.002 | 0.31 ± 0.002 | 64.6 ± 0.1 |
| No Age Embedding | 84.6 ± 0.1 | 52.3 ± 0.2 | 89.1 ± 0.1 | 70.7 ± 0.1 | 39.4 ± 0.2 | 93.2 ± 0.1 | 0.37 ± 0.002 | 0.36 ± 0.002 | 68.1 ± 0.1 |
| No Positional Encoding | 85.3 ± 0.1 | 50.1 ± 0.3 | 90.5 ± 0.1 | 70.3 ± 0.1 | 41.9 ± 0.2 | 93.02 ± 0.1 | 0.38 ± 0.002 | 0.38 ± 0.002 | 68.7 ± 0.1 |
| No Age & Positional Encoding | 84.3 ± 0.1 | 61.4 ± 0.3 | 83.9 ± 0.1 | 72.6 ± 0.1 | 34.3 ± 0.2 | 94.1 ± 0.1 | 0.36 ± 0.002 | 0.34 ± 0.002 | 66.3 ± 0.1 |
| No Disease & Drug Embedding | 76.1 ± 0.1 | 29.98 ± 0.3 | 93.3 ± 0.1 | 61.6 ± 0.1 | 37.9 ± 0.4 | 90.2 ± 0.1 | 0.26 ± 0.003 | 0.26 ± 0.003 | 62.7 ± 0.2 |

## 4. Discussion

In this study we have shown that routinely collected data in primary care health on basic demographic variables, diagnoses and drugs prescribed harbours information to predict the risk of developing CRC. The model ExCaPT, based on the encoder of a transformer, achieved high accuracy, useful to stratify patients that should be refered for CRC screening. The age groups below 50 years of age and after 70 years are especially interesting, because they are not invited to participate in screening in current guidelines.

We compared the predictive accuracy of ExCaPT with a baseline model for sequence data. We selected LSTM, that has been shown to perform well when sequences are not very long, as in our setting. ExCaPT, with a transformer archietcture, significantly outperformed LSTM in all evaluation metrics (Table 6). These improvements suggest that ExCaPT not only achieves higher overall accuracy but also maintains a better balance between sensitivity and specificty, leading to more reliable and consistent predictions.

Looking at the confusion matrix (Figure 4a), the model had good PPV for CRC, 34% for the 12% CRC prevalence in the test dataset. The estimated PPV value was 10% for a CRC prevalence of 3%, similar to the 5 − 10% of the FIT if CRC screening [22]. Our dataset did not have reliable data on adenoma occurrence, the precancerous lesion of CRC, and we could not use it as outcome. Regarding sensitivity, 68% is in a similar range as the FIT, but there is room for improvement. Using a lower cutoff could increase sensitivity, at the cost of higher false positive predictions, and unnecesary colonocopies performed, which are expensive and with some risk of adverse effects. Further evaluation on external datasets and under different conditions would be beneficial to better assess the model’s generalizability and real-world utility.

In this study we did not have information on actual participation in screening to take this variable into account. Screening participation is low in the Catalan population (∼45%). The information from this predictive model could be an element to ecourage participation for reluctant subjects with high predicted risk.

The stratified evaluation, shown in Table 6 highlights several patterns in model performance. In terms of age stratification, the model achieves its highest values across all metrics in the oldest age group (*>*70 years), suggesting that disease patterns in older patients are more distinctive and easier to classify. This age group also has higher CRC prevalence, and all combined makes the prediction more useful. The age group below 50 is of high interest, given the recent concern of a worldwide rising incidence [23]. Though sensitivity to detect early onset CRC is similar than average, the specificity is lower, which combined with low CRC prevalence (10%) in the test dataset results in a PPV of 19%.

When restricting by disease sequence length for patients under 50, performance generally declined as the minimum required length increased, indicating that filtering for longer sequences reduces sample diversity and weakens generalization. Sex-based evaluation showed only marginal differences, with female patients performing slightly better across most metrics, suggesting that the model remains largely fair with respect to sex.

The ablation study in Table 8 demonstrates the contribution of each model component. The *Full Model* achieved the best performance (Balanced Accuracy 75.1%, ROC-AUC 85.9%). Using only the class token (*Only Class Token Enabled* ) caused a major drop (Balanced Accuracy 55.6%, ROC-AUC 61.5%), while removing the class token (*No Class Token*) partially recovered performance (Balanced Accuracy 68.2%, ROC-AUC 84.5%), indicating that mean pooling of token embeddings effectively aggregates sequence information.

Disease and drug embeddings were the most critical features; disabling them (*No Disease & Drug Embedding* ) strongly reduced balanced accuracy (61.6%) and sensitivity (29.98%). Age embedding and positional encoding also contributed meaningfully, particularly to sensitivity, whereas sex and smoking embeddings had minimal impact. Overall, these results show that the model relies primarily on sequence-specific and disease/drug information, with demographic features playing a secondary role.

Regarding explainability, Figure 6a shows that most annotated drugs favor Non-CRC predictions, which aligns with expectations. Some diseases are more widely distributed and appear in both prediction classes. Figure 7 shows that true positives and true negatives form distinct clusters, whereas false positives and false negatives overlap in the middle region of those clusters, indicating that the model learned discriminative representations, with some misclassifications. Regarding IG (Figure 8), most annotated diseases favor CRC predictions, while annotated drugs are sparsely distributed across both classes. Overall, the model captures some discriminative patterns, even though attention and IG analyses suggest it does not strictly separate feature types as might be expected. This indicates that misclassifications occur in ambiguous cases and that the model may rely on complex feature interactions rather than simple disease-drug associations.

Our study had some limitations. The study design, as a matched case-control study was convenient to balance the training, but may suffer from some selection bias and may artificially increase the predictive accuracy metrics, mainly the PPV, because the prevalence of CRC is highly increased compared to that in the population. We used diagnostics coded in the electronic health records, which may be incomplete and not always verified. Smoking habit had a large fraction of missing values. Regarding drugs, we used prescription data, which doesn’t assure the drug was actually consumed by the patient, and we did not have information on over the counter drugs that the patient may adquire out of the health system without a presciption.

Based on obtained results, the ExCaPT model can reasonably distinguish between CRC and Non-CRC cases. In terms of interpretability, the attention mechanism within the transformer architecture highlights factors that contribute to both CRC and Non-CRC predictions, while the learned embeddings effectively capture and differentiate key features.

Compared to traditional CRC screening based on FIT, which only stratifies the population regarding age and family history, the addition of ExCaPT may offer advantages, specially to consider candidates for screening at early age that the standard for screening (50 in Spain). For elderly individuals, when screening is no longer indicated because of limited resources, the use of stratification models like ExCaPT could be an atractive option to be studied regarding cost-effectiveness.

## Data Availability

The data was provided by the Healthcare Quality and Evaluation Agency of Catalonia (AQuAS) and is available under request to this institution.

## Acknowledgments

We thank CERCA Programme, Generalitat de Catalunya for institutional support.

## 5. Funding

This study has been funded by Instituto de Salud Carlos III (ISCIII), “Programa FORTALECE del Ministerio de Ciencia e Innovación”, through the project number FORT23/00032 and D.I. received a Sara Borrell fellowship CD24/00094 (FIS24031). A.M. received a Juan de la Cierva fellowship JDC2023-052616-I from the Ministerio de Ciencia, Innovación y Universidades, Spain. Spanish Science and Innovation Ministry grant "Transformación Energética y Digital" TED2021-132025B-C41. TRANSCAN3 project TANGERINE (TRANSCAN2021-071 AC22/00021) funded by ISCIII and Fundación Científica de la Asociación Española contra el Cancer. DTS project DeepRDT (DTDTS22/0007) funded by ISCIII.

## 6. Author contributions

**Conceptualization:** Victor Moreno

**Data curation:** Ditsuhi Iskandaryan, Ferran Moratalla-Navarro, Alba Magallon-Baro, Lois Riobó-Mayo, Tomas Salas, Miguel Socolovsky, Victor Moreno

**Formal analysis:** Ditsuhi Iskandaryan, Ferran Moratalla-Navarro, Alba Magallon-Baro, Lois Riobó-Mayo, Miguel Socolovsky, Victor Moreno

**Funding acquisition:** Ditsuhi Iskandaryan, Alba Magallon-Baro, Lois Riobó-Mayo, Victor Moreno

**Invstigation:** Ditsuhi Iskandaryan

**Methodology:** Ditsuhi Iskandaryan, Ferran Moratalla-Navarro, Victor Moreno

**Supervision:** Victor Moreno

**Resources:** Tomas Salas

**Visualization:** Ditsuhi Iskandaryan, Ferran Moratalla-Navarro, Alba Magallon-Baro, Lois Riobó-Mayo, Miguel Socolovsky, Victor Moreno

**Writing—original draft:** Ditsuhi Iskandaryan

**Writing—review editing:** Ditsuhi Iskandaryan, Ferran Moratalla-Navarro, Alba Magallon-Baro, Lois Riobó-Mayo, Tomas Salas, Miguel So-colovsky, Victor Moreno

## 7. Competing interests

The authors declare no competing interests.

## Notes

### Competing Interest Statement

The authors have declared no competing interest.

### Funding Statement

Yes

### Author Declarations

The Bellvitge Hospital Ethics Committee approved the study protocol (number PR320/18) waiving the need to obtain informed consent, given the large population analyzed and the anonymization procedure used.

